# Linking Modifiable Risk Factors to Vascular and Neurodegenerative Brain Changes

**DOI:** 10.64898/2026.02.28.26347178

**Authors:** Tamara Khudair, Shima Raeesi, Farooq Kamal, Mahsa Dadar, Cassandra Morrison

## Abstract

**INTRODUCTION:** Dementia reflects vascular and neurodegenerative processes in late life, yet studies often examine risks and outcomes individually. This study tested whether the cumulative burden of risks relates to structural brain pathology and cognition, and whether brain markers mediate these associations.

**METHODS:** Cross-sectional data were drawn from 38,414 older adults in the National Alzheimer’s Coordinating Center database. A composite score summed ten binary risk factors: hypertension, diabetes, hypercholesterolemia, alcohol misuse, smoking, depression, obesity, hearing loss, vision loss, and low education. Outcomes included white matter hyperintensities (WMH), infarcts, hippocampal atrophy, global cognition, cognitive status, delayed recall, and semantic fluency.

**RESULTS:** Higher burden was associated with poorer global cognition, greater clinical severity, worse memory and fluency, and higher odds of WMHs, infarcts, and hippocampal atrophy. Structural equation models identified hippocampal atrophy as the primary mediator, with smaller effects for WMHs and infarcts.

**DISCUSSION:** Findings support multidomain prevention strategies targeting clustered modifiable risks.

## 1. Background

Structural brain changes are among the most frequent and clinically relevant markers of cognitive decline in older adults, including white matter hyperintensities (WMHs), hippocampal atrophy, and cerebral infarcts (1,2). WMHs appear as bright regions on T2-weighted and FLAIR magnetic resonance imaging (MRI) scans (or as hypointensities on T1-weighted MRIs) and reflect chronic ischemia and microvascular damage (3,4,6). They increase with age and are more extensive in individuals with vascular conditions such as hypertension and diabetes (5). WMHs are widely recognized as markers of cerebral small vessel disease and are associated with slower processing speed, weaker executive function, and an increased risk of developing mild cognitive impairment (MCI) and dementia (6–8).

In addition to WMHs, cerebral infarcts, including lacunar infarcts and other small ischemic lesions, reflect focal tissue damage caused by reduced blood flow and are associated with poorer cognition and increased dementia risk. Infarcts frequently co-occur with WMHs, reflecting a broader burden of small vessel disease (9,10). Hippocampal atrophy represents a key neurodegenerative pathway to cognitive decline, with volume loss strongly linked to impaired episodic memory and early declines in Alzheimer’s disease (AD) (11,12). Even relatively small reductions in hippocampal size predict future decline, making it a sensitive indicator of neurodegenerative change (13).

A number of modifiable risk factors increase the likelihood of these brain changes. For example, the 2025 Lancet Commission identified 14 modifiable risk factors including excessive alcohol consumption, smoking, high LDL cholesterol, hypertension, diabetes, depression, hearing loss, vision loss, obesity, low education, physical inactivity, traumatic brain injury, air pollution, and social isolation. These risk factors are common, often co-occur, and contribute to both vascular injury and reduced cognitive resilience in older adults (14).

Despite their frequent co-occurrence, most studies have examined them individually (15–25). Moreover, studies that do consider multiple risk factors often focus on one outcome, such as WMH or a single cognitive domain, limiting understanding of how cumulative risk burden relates to multiple aspects of brain structure and function (26–29). This gap highlights the need for research examining the combined influence of several risk factors across multiple vascular and cognitive outcomes. Given that these exposures frequently cluster, assessing their total burden and progression can clarify how multiple risks contribute to WMH accumulation, hippocampal atrophy, infarcts, and cognitive decline.

The primary objective of this study is to address these gaps by testing whether a higher burden of documented or currently present modifiable risk factors shows a dose-response pattern with key markers of vascular and cognitive health. Specifically, the study evaluates whether cognitive and brain outcomes progressively worsen as the number of current risk factors increase. The analysis draws on data from the National Alzheimer’s Coordinating Center (NACC) database (30), which includes comprehensive clinical, MRI, and neuropsychological assessments from participants across the United States, offering a well-characterized sample of aging adults. Demonstrating a dose-response graded relationship between total risk factor burden and measures such as WMH volume, hippocampal atrophy, infarcts, and cognition would provide important evidence that the accumulation of co-occurring risks meaningfully contributes to brain vulnerability and may inform prevention strategies aimed at preserving cognitive and vascular brain health.

## 2. Methods

### 2.1 Data source and participants

Data were obtained from the NACC database, including the Uniform Data Set Version 3 (UDS v3) and linked MRI variables (31,32). The analytic sample was constructed using a cross-sectional design, selecting a single baseline visit per participant. Baseline was defined as the earliest visit with valid cognitive, MRI, and risk-factor data. Participants aged 55 years and older at baseline were eligible for inclusion.

Race was derived from UDS v3 fields and harmonized into mutually exclusive categories (White, Black, Asian), excluding unknown or other-specification codes with insufficient detail. Cognitive status was calculated using the NACCTMCI and NACCETPR variables (representing the primary diagnosis) as well as Clinical Dementia Rating-Sum of Boxes (CDR-SB) scores, ensuring consistency in cognitive categorization. Participants with CDR-SB of 0 and no neurological disorders based on NACCETPR variable were classified as “NC”, those with a CDR-SB between 0.5 and 4 were classified as “MCI,” and those with CDR-SB > 4.5 were classified as “AD”. Participants with other neurological disorders (Parkinson’s disease, Lewy body dementia, frontotemporal dementia, etc.) or inconsistent CDR-SB scores and diagnostic labels (e.g., individuals with CDR-SB > 4.5 and no diagnosis identified based on NACCETPR) were excluded. Participants with these inconsistencies were excluded according to NACC recommendations (31,32).

### 2.2 Modifiable risk factors and composite burden

Ten modifiable risk factors were derived from UDS v3 health history and examination variables: hypertension, diabetes, hypercholesterolemia (high LDL cholesterol), alcohol misuse, cigarette smoking, depression, obesity, hearing loss, vision loss, and low education. Each factor was coded as a binary indicator (0 = no evidence of the risk factor, 1 = evidence of the risk factor), reflecting any documented history or current presence at the baseline visit, participants without information for all ten factors were excluded from the composite analyses.

Hypertension, diabetes, and hypercholesterolemia were coded using NACC UDS v3 health history variables. Hypertension was classified as present when the HYPERTEN variable indicated a history of high blood pressure and absent when HYPERTEN explicitly indicated no history. Diabetes was classified using the DIABETES variable, with participants identified as having diabetes when a diagnosis was recorded and as not having diabetes when DIABETES indicated no history. Hypercholesterolemia was determined from the HYPERCHO variable, with a recorded diagnosis indicating hypercholesterolemia and an explicit “no history” response indicating its absence.

Alcohol misuse was coded using the NACC UDS v3 ALCOHOL variable. Participants were classified as having alcohol misuse when ALCOHOL indicated problem use associated with clinically significant impairment in work, social, legal, or daily functioning. Those for whom ALCOHOL indicated no such history were classified as having no alcohol misuse. Depression was coded as present when current or prior depressive episodes or related diagnoses were recorded (DEP, DEPD, DEPOTHR), and absent when all depression fields indicated no history.

Smoking status was coded as a binary variable indicating ever smoking versus never smoking using NACC UDS v3 smoking history variables. Participants were classified as ever smokers if any evidence of cigarette use was present, including nonzero packs per day (PACKSPER), nonzero years smoked (SMOKYRS), endorsement of smoking within the past 30 days (TOBAC30), or documentation of having previously quit smoking (QUITSMOK). Participants were classified as never smokers when PACKSPER indicated zero use and no smoking history variables suggested prior cigarette use.

Hearing and vision status was derived from NACC UDS v3 sensory variables. Hearing classification used HEARING (hearing without aids), HEARAID (hearing-aid use), and HEARWAID (hearing with aids). Participants were classified as having hearing loss when hearing was rated as not normal and remained impaired even with hearing aids. They were classified as having normal hearing when hearing was normal without aids or when impairment was fully corrected with aids. Vision classification used VISION (vision without correction), VISCORR (use of corrective lenses), and VISWCORR (vision with correction). Participants were classified as having vision loss when vision was impaired without correction and remained impaired even with lenses, and as having normal vision when vision was normal unaided or fully corrected.

Obesity status was based on body mass index (BMI) from the NACCBMI variable. Participants with BMI ≥ 30 were classified as obese, and those with BMI < 30 were classified as non-obese (33). Education was coded using the EDUC variable. Participants with fewer than 12 years of formal schooling were classified as having low education, while those with 12 or more years were classified as having higher education.

A composite risk-burden index was then created by summing the ten binary risk-factor indicators for each participant. This composite ranged from 0 to 10 and captured the total number of modifiable risk factors present. Participants with missing values for all ten risk factors were excluded from composite analyses.

### 2.3 Cognitive and clinical outcomes

Cognitive outcomes were drawn from UDS v3 neuropsychological measures and global ratings. Global cognitive status was indexed using the NACC variable COGSTAT, which reflects clinician judgment of overall cognitive functioning based on neuropsychological test performance, with higher scores indicating worse overall cognitive status. Global clinical severity was assessed using the CDR-SB, with higher scores indicating greater dementia-related impairment in cognition and daily functioning.

Two domain-specific cognitive measures were selected to capture complementary aspects of late-life cognition. Verbal episodic memory was assessed using the Craft Story delayed recall score (CRAFTDVR), which reflects delayed recall of a narrated story and is sensitive to hippocampal and medial temporal lobe integrity (34,35). Semantic fluency was measured using Category Fluency - Animals (ANIMALS), requiring participants to generate animal names within a fixed time and indexing both semantic knowledge and executive search processes (36–38). Higher scores on CRAFTDVR and ANIMALS indicate better performance.

### 2.4 Vascular and neurodegenerative MRI outcomes

Vascular and neurodegenerative brain outcomes were derived from NACC imaging variables linked to the UDS v3 baseline visit. White matter hyperintensities (WMHs) were captured using two binary MRI indicators: IMAGMWMH, which denotes moderate WMH burden (CHS score 5-6), and IMAGEWMH, which denotes extensive WMH burden (CHS score 7-8). A combined WMH indicator was created by coding WMH as present when either IMAGMWMH or IMAGEWMH was coded “yes”, and as absent when both variables were coded “no”.

Cerebral infarcts were measured using NACC MRI variables indexing large vessel and lacunar infarcts. Large vessel infarcts were taken from IMAGLINF, and lacunar infarcts from IMAGLAC, each coded as 0 = no and 1 = yes. A composite infarct variable was created and coded as present when either IMAGLINF or IMAGLAC indicated an infarct, and as absent when both variables indicated no infarct.

Hippocampal atrophy was indexed with the HIPPATR variable, which reflects clinician-rated hippocampal atrophy based on MRI according to each center’s local imaging protocols. Responses indicating definite atrophy were classified as hippocampal atrophy present, whereas responses indicating no atrophy were classified as absent.

### 2.5 Statistical Analysis

Descriptive statistics were used to summarize demographic, clinical, cognitive, and MRI characteristics at the baseline visit and are presented in Table 1.

**Table 1.** Demographic, Clinical, and Neuroimaging Characteristics for Study Participants.

| Characteristic | Total (n = 38,414) |
| --- | --- |
| Age, mean (SD) | 72.97 (8.68) |
| Female, n (%) | 22,494 (58.6%) |
| White, n (%) | 31,230 (81.3%) |
| Black, n (%) | 6,120 (15.9%) |
| Asian, n (%) | 1,064 (2.8%) |
| Education, years, mean (SD) | 15.38 (3.27) |
| NC, n (%) | 16,982 (44.1%) |
| MCI, n (%) | 13,599 (35.4%) |
| AD, n (%) | 7,833 (20.4%) |
| Diabetes, n (%) | 5,093 (13.3%) |
| Hypertension, n (%) | 19,477 (50.9%) |
| Hypercholesterolemia, n (%) | 19,876 (52.3%) |
| Alcohol misuse, n (%) | 1,796 (4.7%) |
| Smoking history, n (%) | 16,142 (42.8%) |
| Depression, n (%) | 15,150 (41.3%) |
| Obesity, n (%) | 8,303 (24.0%) |
| Hearing loss, n (%) | 572 (1.7%) |
| Vision loss, n (%) | 1,261 (3.9%) |
| Low education, n (%) | 2,315 (6.1%) |
| WMH present, n (%) | 1,054 (15.6%) |
| Infarcts present, n (%) | 596 (8.7%) |
| Hippocampal atrophy, n (%) | 4,273 (41.4%) |

NC = Cognitively Normal. MCI = Mild Cognitive Impairment. AD = Alzheimer's disease. SD = Standard Deviation. WMH = White matter hyperintensity. Values are presented as total number and percentage or mean and standard deviation. Diabetes, hypertension, hypercholesterolemia, alcohol misuse, smoking history, obesity, hearing loss, vision loss, low education were reported as 1 (has the condition) or 0 (does not have the condition).

To examine whether a higher number of currently present risk factors was related to poorer cognitive or brain outcomes, regression models were completed for each outcome. Linear regression was used for continuous cognitive measures, including global cognitive status, global clinical severity, delayed recall, and semantic fluency. Logistic regression was used for binary MRI outcomes, including white matter hyperintensity presence, infarcts, and hippocampal atrophy. In these models, the composite count of currently present risk factors was the main predictor. All models adjusted for age, sex, and race. Education was not included as a covariate because low education was incorporated directly into the composite risk index.

In addition to the composite score, each of the ten individual risk factors were examined separately. For cognitive outcomes, each factor was entered into a separate linear regression model. For MRI outcomes, each factor was examined using logistic regression. These models used the same covariates as the composite analyses. The *p*-values are reported as raw values with significance determined using false discovery rate (FDR) correction applied within each outcome (39).

For linear regression models, standardized coefficients were used to describe adjusted associations. For logistic models, odds ratios were used to summarize effect sizes. These analyses allowed us to evaluate both the combined effect of multiple co-occurring risks and the specific contribution of individual risk factors.

For analysis, all continuous variables, such as COGSTAT, CDRSUM, delayed recall and semantic fluency, were standardized to z scores. This method allowed coefficients to be interpreted as standardized mean differences in each outcome per unit increase in the composite risk index, controlling for covariates.

The interdependent relationships between risk factors, brain pathology, and cognitive outcomes were further investigated using structural equation modeling (SEM) with the lavaan package (version 0.6-19) in R. Analyses were conducted in the subset of participants with complete MRI and cognitive data to examine direct and indirect pathways from individual risk factors to cognitive outcomes through cerebrovascular and neurodegenerative markers. The model specified a three-level structure, with ten modifiable risk factors as predictor variables, three MRI-based markers (WMHs, cerebral infarcts, and hippocampal atrophy) as parallel mediators, and four cognitive outcomes (global cognitive status, CDR-SB, delayed recall, and semantic fluency) as downstream variables. Risk factors were specified to predict each brain mediator, and brain mediators were specified to predict each cognitive outcome. Direct paths from risk factors to cognitive outcomes were simultaneously included to estimate associations not accounted for by the brain mediators. Correlations among risk factors and among brain mediators were freely estimated to account for shared etiological and pathological processes; these covariances are not reported, as they were not central to the study aims. Age, sex, and race were included as covariates for all mediators and cognitive outcomes, consistent with the primary regression analyses. Because the brain mediators were binary variables, models were estimated using diagonally weighted least squares with the weighted least squares mean- and variance-adjusted estimator (WLSMV). Brain measures were specified as ordered categorical endogenous variables. Models were estimated using the theta parameterization with mean structures included. This estimation approach provides robust standard errors and appropriate model test statistics for models including categorical mediators and continuous outcomes.

Model fit was evaluated using the chi-square statistic, comparative fit index (CFI), Tucker–Lewis index (TLI), root mean square error of approximation (RMSEA), and standardized root mean square residual (SRMR). Because all possible direct and indirect paths were specified to examine the full network of associations, the model was saturated (degrees of freedom = 0), therefore, global fit indices were interpreted with caution, and inference focused primarily on individual path estimates. Standardized path coefficients (Std.all) are reported to facilitate comparison of effect sizes across pathways. Indirect effects were computed as the product of the risk–brain (a-path) and brain–cognition (b-path) coefficients, and total effects were calculated as the sum of direct and indirect effects. Statistical significance was evaluated at a two-sided α level of 0.05.

## 3. Results

### 3.1 Demographics and Clinical Data

Table 1 summarizes the demographic, clinical, and neuroimaging characteristics of the study sample. The cohort included 38,414 older adults, with a mean age of 72.97 ± 8.68 years, and 58.6% of participants were female. Most individuals identified as White (81.3%), with smaller proportions identifying as Black (15.9%) or Asian (2.8%). Mean educational attainment was 15.38 ± 3.27 years, and 6.1% of participants had fewer than 12 years of formal education.

Modifiable vascular and lifestyle risk factors were common in the sample. Hypertension (50.9%) and hypercholesterolemia (52.3%) were the most prevalent conditions, followed by a history of cigarette smoking (42.8%). Obesity was present in 24.0% of participants, while diabetes was reported in 13.3%. Alcohol misuse was relatively infrequent, affecting 4.7% of the cohort. Sensory impairments were uncommon overall, with hearing loss present in 1.7% and vision loss in 3.9% of participants. Neuroimaging markers of brain pathology were also prevalent. White matter hyperintensities were present in 15.6% of participants, cerebral infarcts in 8.7%, and hippocampal atrophy in 41.4%. Cognitive diagnoses at baseline included cognitively normal status in 44.1% of participants, mild cognitive impairment in 35.4%, and Alzheimer’s dementia in 20.4%.

### 3.2 Composite Risk Factor Profiles and Cognition

Higher composite risk was consistently associated with poorer cognitive functioning across multiple domains. Each one-unit increase in the composite risk score was linked to worse global cognitive status (COGSTAT: β = 0.05, *t* = 15.35*, p* < .001) and greater global clinical severity (CDR-SB: β = 0.03, *t* = 8.15, *p* < .001). Neuropsychological performance also declined with increasing risk burden: participants scored lower on episodic memory (CRAFT β = -0.03, *t* = -5.36*, p* < .001) and semantic fluency (ANIMALS β = -0.05, *t* = -15.45*, p* < .001).

### 3.3 Composite Risk Factor Profile and Brain Health Outcomes

Risk burden was strongly linked to markers of cerebrovascular and neurodegenerative injury. Each additional point increase in composite risk score increased the odds of hippocampal atrophy by approximately 5% (OR = 1.05, *p* < .001). Associations with cerebrovascular pathology were even greater: the odds of having moderate or extensive WMH burden increased by 19% (OR = 1.19, *p* < .001), and the odds of cerebral infarcts increased by 18% (OR = 1.18, *p* < .001) for every additional risk factor endorsed.

### 3.4 Individual Risk Factors and Cognitive and Brain Health Outcomes

Table 4 provides the estimates and *p*-values for each risk factor across all vascular and cognitive outcomes. Across the individual risk factors, several clear patterns emerged in their associations with vascular brain injury and cognitive outcomes. The risk factors most strongly associated with cerebrovascular injury were hypertension, hypercholesterolemia, diabetes, smoking history, and alcohol misuse. Hypertension showed the largest vascular effects, with higher odds of both moderate or extensive WMHs (OR = 1.68, *p* < .001) and infarcts (OR = 1.65, *p* < .001). Hypercholesterolemia and diabetes showed similar patterns, as both were associated with higher odds of WMHs (hypercholesterolemia: OR = 1.33, *p* < .001; diabetes: OR = 1.33, *p* < .001) and infarcts (hypercholesterolemia: OR = 1.20, *p* = 0.006; diabetes: OR = 1.47, *p* < .001). Alcohol misuse was also linked to increased odds of WMHs (OR = 1.43, *p* = 0.004) and infarcts (OR = 1.69, *p* < .001), while smoking history showed a smaller effect on WMHs (OR = 1.16, *p* = 0.006) and was not related to the presence of infarcts. Low education was also related to higher odds of infarcts (OR = 1.57, *p* = 0.005). In contrast, hippocampal atrophy was most strongly associated with depression (OR = 1.42, *p* < .001) and vision loss (OR = 1.39, *p* = 0.005), while obesity showed the opposite pattern, with lower odds of hippocampal atrophy (OR = 0.76, *p* < .001).

Cognitive outcomes showed a different profile of associations. Depression and low education demonstrated the broadest and most consistent links to poorer cognitive performance. Depression was associated with worse cognitive status (β = 0.21, *p* < .001), worse clinical severity (β = 0.35, *p* < .001), and poorer performance in both delayed recall (β = -0.24, *p* < .001) and semantic fluency (β = -0.26, *p* < .001). Low education showed a similar pattern and even larger effects, with worse cognitive status (β = 0.25, *p* < .001), substantially higher clinical severity (β = 0.60, *p* < .001), poorer delayed recall (β = -0.57, *p* < .001), and weaker semantic fluency (β = -0.58, *p* < .001).

Several vascular risk factors were associated with lower CDR-SB scores, including hypercholesterolemia (β = -0.06, *p* < .001), hypertension (β = -0.02, *p* = 0.031), obesity (β = -0.12, *p* < .001), and smoking history (β = -0.08, *p* < .001), indicating lower overall clinical severity despite their adverse vascular profiles. These associations contrast with the cognitive test results for obesity and smoking, where both showed small but statistically significant positive coefficients for delayed recall (obesity β = 0.14, *p* < .001; smoking β = 0.06, *p* < .001) and semantic fluency (obesity β = 0.06, *p* < .001; smoking β = 0.04, *p* < .001).

**Table 2.**
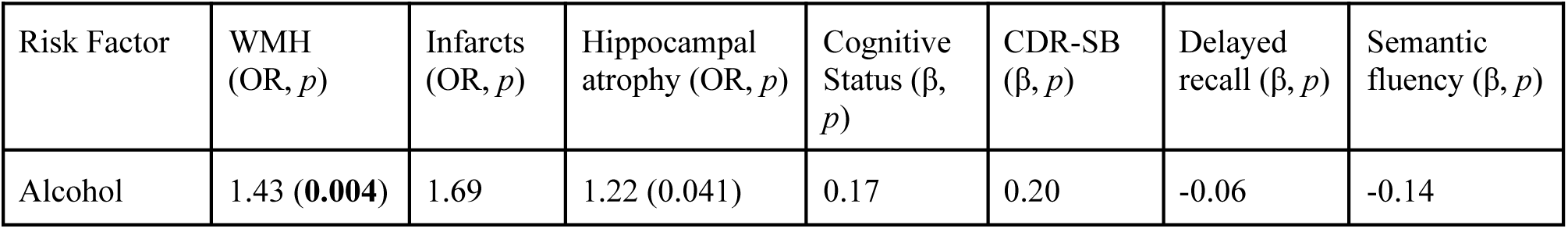
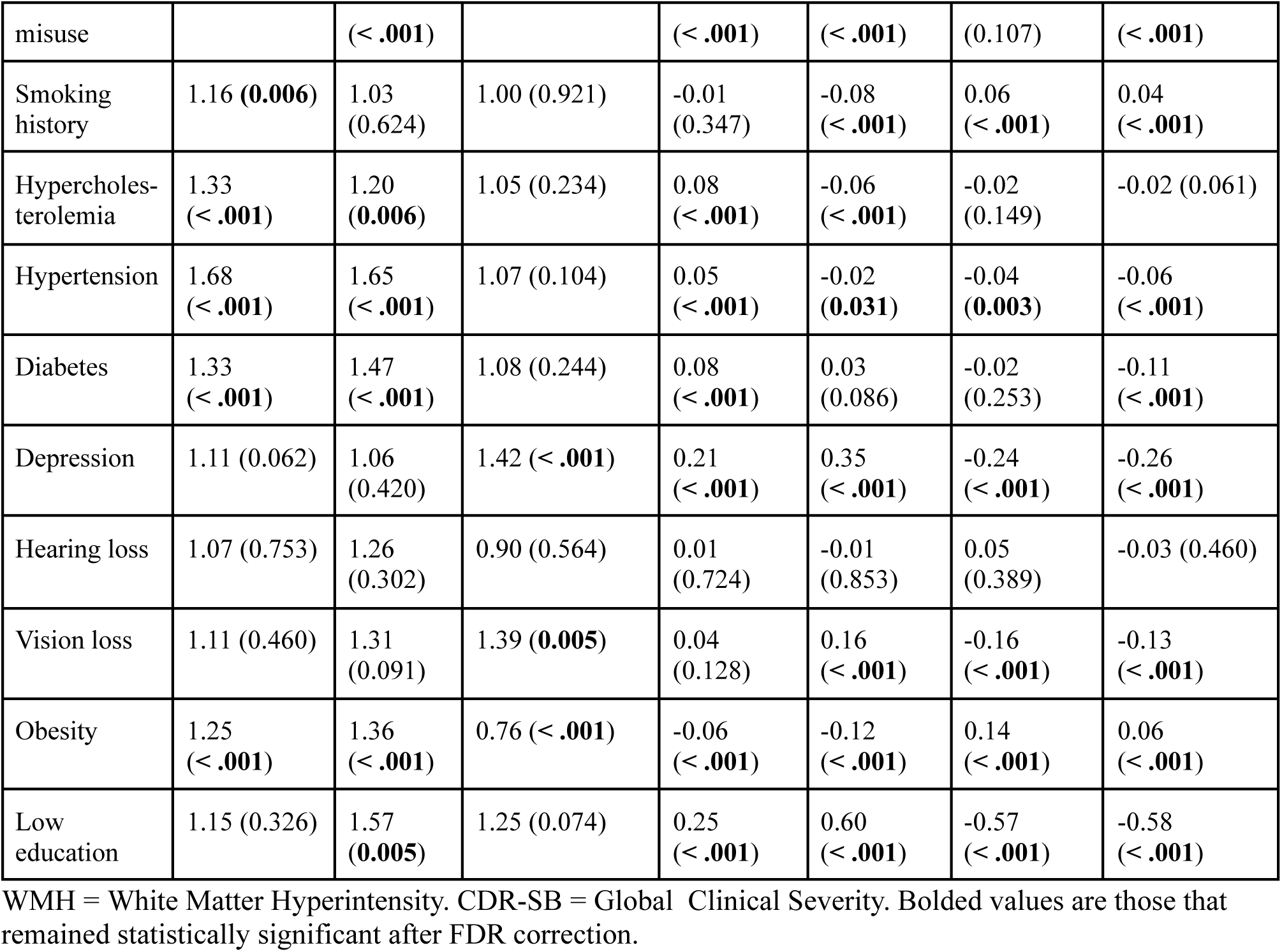
Associations Between Individual Risk Factors and All Brain & Cognitive Outcomes.

### 3.5 Structural Equation Modeling of Risk Factor → Brain Pathology → Cognition Pathways

Structural equation modeling evaluated simultaneous direct and indirect pathways from individual risk factors to cognition through WMHs, infarcts, and hippocampal atrophy. Because the SEM was saturated (df = 0), global fit indices are not informative; interpretation focused on path estimates and indirect/total effects (Tables 3-5).

**Table 3.** SEM Model Results: Direct Associations Between Risk Factors and Brain (a-paths) and Cognitive Outcomes (c′-paths)

|  | a-paths |  |  | c'-paths |  |  |  |
| --- | --- | --- | --- | --- | --- | --- | --- |
| Risk Factor | WMH<br>(Std.all, $p$) | Infarcts<br>(Std.all, $p$) | Hippocampal atrophy<br>(Std.all, $p$) | Cognitive Status<br>(Std.all, $p$) | CDR-SB<br>(Std.all, $p$) | Delayed recall<br>(Std.all, $p$) | Semantic fluency<br>(Std.all, $p$) |
| Alcohol misuse | 0.04 (0.06) | 0.05 (**0.04**) | -0.01 (0.59) | 0.00 (0.85) | 0.00 (0.80) | 0.01 (0.42) | 0.02 (0.14) |
| Smoking history | 0.00 (0.96) | -0.04 (0.17) | 0.00 (0.91) | 0.00 (0.78) | -0.06 (**<.001**) | 0.03 (0.06) | 0.02 (0.10) |
| Hypercholesterolemia | 0.03 (0.22) | 0.05 (0.09) | 0.01 (0.46) | 0.04 (**0.01**) | -0.01 (0.50) | -0.02 (0.26) | -0.01 (0.47) |
| Hypertension | 0.14 (**<.001**) | 0.11 (**<.001**) | 0.04 (0.06) | -0.01 (0.36) | 0.01 (0.69) | -0.02 (0.12) | -0.02 (0.27) |
| Diabetes | 0.01 (0.75) | 0.02 (0.41) | 0.02 (0.36) | 0.00 (0.83) | -0.01 (0.37) | 0.02 (0.26) | 0.00 (0.83) |
| Depression | 0.01 (0.63) | 0.03 (0.24) | 0.09 (**<.001**) | 0.04 (**0.02**) | 0.09 (**<.001**) | -0.05 (**0.002**) | -0.07 (**<.001**) |
| Hearing loss | 0.01 (0.47) | 0.04 (0.09) | 0.03 (0.13) | 0.00 (0.55) | 0.00 (0.59) | -0.01 (0.05) | 0.00 (0.88) |
| Vision loss | 0.00 (0.83) | -0.03 (0.33) | 0.02 (0.26) | 0.01 (0.44) | 0.00 (0.80) | 0.00 (0.92) | 0.00 (0.62) |
| Obesity | 0.02 (0.41) | 0.00 (0.92) | -0.08 (**<.001**) | 0.02 (0.20) | -0.03 (0.07) | 0.05 (**0.003**) | 0.00 (0.95) |
| Low education | 0.01 (0.54) | 0.01 (0.68) | 0.00 (0.82) | 0.04 (0.06) | 0.02 (0.24) | -0.05 (**0.01**) | -0.07 (**<.001**) |

WMH = White Matter Hyperintensity. CDR-SB = Global Clinical Severity. Bolded values represent statistically significant $p$ -values.

**Table 4.** SEM Model Results: Direct Associations Between Brain Measures and Cognitive Outcomes (b-paths)

|  | **b-paths** |  |  |  |
| --- | --- | --- | --- | --- |
| Brain Measures | Cognitive Status<br>(Std.all, $p$) | CDR-SB<br>(Std.all, $p$) | Delayed recall<br>(Std.all, $p$) | Semantic fluency<br>(Std.all, $p$) |
| WMH | 0.07 (0.05) | 0.02 (0.51) | 0.02 (0.56) | -0.06 (0.08) |
| Infarcts | 0.07 (0.06) | -0.03 (0.38) | -0.02 (0.55) | 0.00 (0.96) |
| Hippocampal atrophy | **0.27 (<.001)** | **0.55 (<.001)** | **-0.49 (<.001)** | **-0.38 (<.001)** |

WMH = White Matter Hyperintensity. CDR-SB = Global Clinical Severity. Bolded values represent statistically significant $p$ -values.

**Table 5.** SEM Model Results: Indirect Associations Between Risk Factors and Cognitive Outcomes (c-paths) and Total Effects.

|  | **c-paths** |  |  |  | **Total Effects** |  |  |  |
| --- | --- | --- | --- | --- | --- | --- | --- | --- |
| Risk Factor | Cognitive Status<br>(Std.all,<br>$p$) | CDR-SB<br>(Std.all,<br>$p$) | Delayed recall<br>(Std.all,<br>$p$) | Semantic fluency<br>(Std.all,<br>$p$) | Cognitive Status<br>(Std.all,<br>$p$) | CDR-SB<br>(Std.all,<br>$p$) | Delayed recall<br>(Std.all,<br>$p$) | Semantic fluency<br>(Std.all,<br>$p$) |
| Alcohol misuse | 0.00<br>(0.52) | -0.01<br>(0.55) | 0.00<br>(0.62) | 0.00<br>(0.88) | 0.01<br>(0.67) | -0.01<br>(0.53) | 0.01<br>(0.29) | 0.02<br>(0.13) |
| Smoking history | 0.00<br>(0.62) | 0.00<br>(0.99) | 0.00<br>(0.84) | 0.00<br>(0.91) | 0.00<br>(0.93) | -0.06<br>(**<.001**) | 0.03<br>(0.05) | 0.03<br>(0.10) |
| Hypercholesterolemia | 0.01<br>(0.15) | 0.01<br>(0.52) | -0.01<br>(0.44) | -0.01<br>(0.36) | 0.05<br>(**0.001**) | 0.00<br>(0.82) | -0.02<br>(0.12) | -0.02<br>(0.24) |
| Hypertension | 0.03<br>(**<.001**) | 0.02<br>(0.08) | -0.02<br>(0.08) | -0.02<br>(**0.01**) | 0.01<br>(0.47) | 0.03<br>(0.09) | -0.04<br>(**0.009**) | -0.04<br>(**0.01**) |
| Diabetes | 0.01<br>(0.27) | 0.01<br>(0.39) | -0.01<br>(0.34) | -0.01<br>(0.35) | 0.01<br>(0.54) | 0.00<br>(0.76) | 0.01<br>(0.59) | -0.01<br>(0.53) |
| Depression | 0.03<br>**(<.001)** | 0.05<br>**(<.001)** | -0.05<br>**(<.001)** | -0.04<br>**(<.001)** | 0.06<br>**(<.001)** | 0.14<br>**(<.001)** | -0.09<br>**(<.001)** | -0.10<br>**(<.001)** |
| Hearing loss | 0.00<br>(0.48) | -0.02<br>(0.11) | 0.01<br>(0.14) | 0.01<br>(0.19) | -0.01<br>(0.40) | -0.02<br>(0.13) | 0.00<br>(0.91) | 0.01<br>(0.67) |
| Vision loss | 0.00<br>(0.52) | 0.01<br>(0.23) | -0.01<br>(0.30) | -0.01<br>(0.28) | 0.02<br>(0.30) | 0.01<br>(0.63) | -0.01<br>(0.52) | -0.01<br>(0.33) |
| Obesity | -0.02<br>**(0.003)** | -0.04<br>**(<.001)** | 0.04<br>**(<.001)** | 0.03<br>**(<.001)** | 0.00<br>(0.91) | -0.07<br>**(<.001)** | 0.08<br>**(<.001)** | 0.03<br>(0.07) |
| Low education | 0.00<br>(0.66) | 0.00<br>(0.83) | 0.00<br>(0.82) | 0.00<br>(0.76) | 0.04<br>(0.05) | 0.02<br>(0.17) | -0.05<br>**(0.004)** | -0.07<br>**(<.001)** |

CDR-SB = Global Clinical Severity. Bolded values represent statistically significant *p*-values.

Direct associations between risk factors and brain measures (SEM a-paths, Table 3): Hypertension was directly associated with WMHs (Std.all = 0.14, *p* < .001) and infarcts (Std.all = 0.11, *p* < .001). Depression was associated with greater hippocampal atrophy (Std.all = 0.09, *p* < .001), whereas obesity was associated with lower hippocampal atrophy (Std.all = -0.08, *p* < .001). Alcohol misuse showed a direct association with infarcts (Std.all = 0.05, *p* = 0.04), while other risk factors were not significantly associated with brain markers.

Direct associations between brain measures and and cognitive outcomes (SEM b-paths): Hippocampal atrophy showed strong associations with all cognitive outcomes (Cognitive status: Std.all = 0.27; CDR-SB: 0.55; Delayed recall: -0.49; Semantic fluency: -0.38; all *p* < .001), while WMHs and infarcts were not independently associated with cognition in the fully adjusted model (Table 4).

Direct associations between risk factors and and cognitive outcomes (SEM c′-paths): Among direct effects of risk factors on cognition (Table 3), depression was associated with worse cognitive status, greater clinical severity (CDR-SB), poorer delayed recall, and lower semantic fluency (all *p* < .01). Smoking history was associated with lower CDR_SB (Std.all = -0.06, *p* < .001), obesity with better delayed recall (Std.all = 0.05, *p* = 0.003), and low education with poorer delayed recall and semantic fluency (both *p* < .01).

Indirect effects (SEM c-paths, Table 5, Fig. 1): Total indirect effects (combined across the three mediators) were observed for depression (all outcomes *p* < .001) and obesity (all outcomes *p* < .001), with smaller indirect effects for hypertension on cognitive status (Std.all = 0.03, *p* < .001) and semantic fluency (Std.all = -0.02, p = 0.01). No significant total indirect effects were observed for the remaining risk factors.

**Figure 1.**
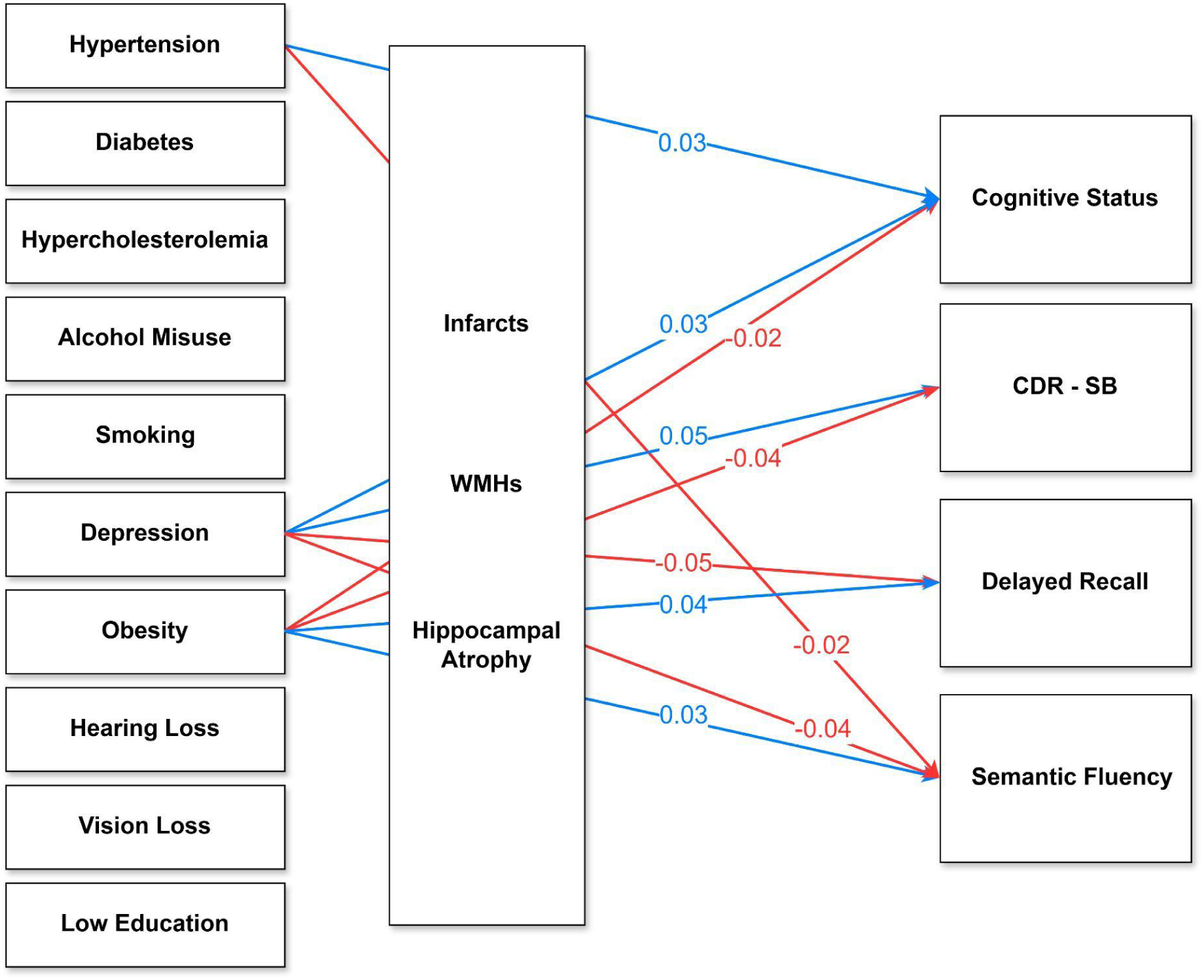
Total indirect (mediated) effects of modifiable risk factors on cognitive outcomes via brain pathology. Figure 1 depicts the structural equation model (SEM) linking ten modifiable risk factors (left) to MRI markers of brain pathology (middle: infarcts, WMHs, hippocampal atrophy) and downstream cognitive outcomes (right: cognitive status (COGSTAT), CDR-SB, delayed recall, semantic fluency). Numbers on arrows represent standardized total indirect effects (Std.all), reflecting the combined mediated association from each risk factor to each cognitive outcome through the three brain markers. Only statistically significant paths are displayed. Blue arrows indicate positive associations and red arrows indicate negative associations. The SEM was estimated using WLSMV with binary brain mediators modeled as ordered categorical variables and adjusted for age, sex, and race for all mediators and outcomes. All cognitive outcomes were standardized (z-scores). Higher values of COGSTAT and CDR-SB indicate worse cognitive functioning and greater clinical severity, whereas higher delayed recall and semantic fluency scores reflect better cognitive performance.

Total effects (direct + indirect associations) were strongest and most consistent for depression (all outcomes *p* < .001). Obesity was associated with lower CDR-SB and higher delayed recall (both *p* < .001). Smoking history was associated with lower CDR-SB (Std.all = -0.06, *p* < .001), and hypertension and low education showed negative total effects on delayed recall and semantic fluency (all *p* < .02). Hypercholesterolemia was also associated with poorer global cognitive status (Std.all = 0.05, *p* = 0.001). Some inverse associations may reflect residual confounding, survival bias, or reverse causation rather than clinically protective effects (Tables 4, S1, Fig. 2).

**Figure 2.**
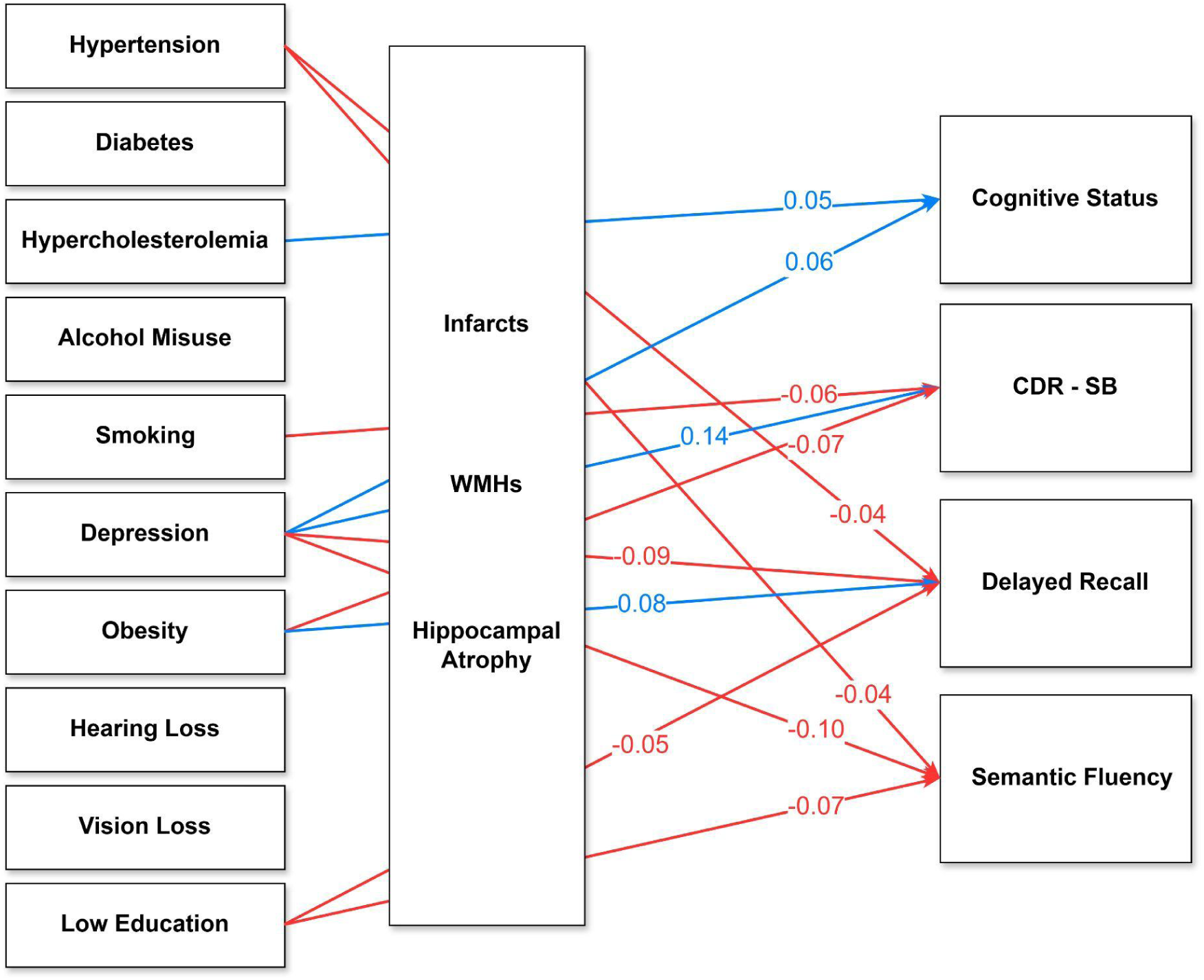
Structural equation model showing total effects of risk factors on cognitive outcomes. Figure 2 shows the same SEM framework as Figure 1, with numbers on arrows representing standardized total effects (Std.all; direct + indirect) from each risk factor to each cognitive outcome. Only statistically significant paths are displayed. Blue arrows indicate positive effects and red arrows indicate negative effects. The SEM was estimated using WLSMV with binary brain mediators modeled as ordered categorical variables and adjusted for age, sex, and race for all mediators and outcomes. All cognitive outcomes were standardized (z-scores). Higher values of COGSTAT and CDR-SB indicate worse cognitive functioning and greater clinical severity, whereas higher delayed recall and semantic fluency scores reflect better cognitive performance.

## 4. Discussion

This study examined the cumulative burden of ten modifiable risk factors in relation to vascular brain injury, neurodegenerative markers, and cognitive performance in a large cohort of older adults. The findings demonstrate a robust, incremental pattern in which greater co-occurring risk burden corresponds to a higher likelihood of structural brain injury, including WMH burden, cerebral infarcts, and hippocampal atrophy. These associations extended across cognitive outcomes, with increasing cumulative risk linked to worse global cognitive status, greater clinical severity, and poorer delayed recall and semantic fluency. Together, these results reinforce the concept that modifiable risks act in an additive manner, with their combined presence exerting a stronger influence on late-life brain health than any single factor alone.

These analyses extend prior work by showing that cumulative exposure across vascular, metabolic, sensory, and lifestyle factors influences brain aging in interconnected ways (40–43). Much of the existing literature has focused on single risk factors or narrow outcomes, such as the effects of hypertension on WMHs or the influence of diabetes on cognitive decline (15, 22). Fewer studies have assessed multiple modifiable risks simultaneously across structural and functional domains (26–29). By examining ten established risk factors across neuroimaging and cognitive outcomes, this study provides a more comprehensive view of how cumulative exposure relates to late-life brain integrity. The consistent dose-response pattern suggests that a simple count of current risks may serve as an accessible and clinically meaningful indicator of overall brain health vulnerability.

The individual risk-factor analyses added nuance by identifying which factors showed the strongest associations. Hypertension, hypercholesterolemia, diabetes, smoking, and alcohol misuse were most consistently associated with WMHs and infarcts, aligning with extensive evidence that vascular and metabolic dysfunction are central drivers of small vessel disease (10,44–46). Depression and low education showed the broadest associations with cognitive outcomes, spanning global cognitive status, global clinical severity, episodic memory, and semantic fluency (47). These findings underscore that cognitive vulnerability reflects both vascular pathology as well as psychosocial and experiential factors influencing reserve and resilience (48,49).

Structural equation modeling provided additional insight into pathways linking modifiable risk factors and cognition. In the fully adjusted model, hippocampal atrophy emerged as the brain marker most consistently associated with cognitive outcomes, showing robust associations with all measures (50, 51). In contrast, WMHs and infarcts were not independently associated with cognition after accounting for hippocampal atrophy and covariates, possibly reflecting shared variance across brain markers (52–54) and limited sensitivity of binary imaging indicators. Hypertension showed the expected direct associations with vascular pathology (15, 16) but only modest indirect cognitive effects, suggesting that its cognitive impact may be cumulative or unfold over longer periods than captured cross-sectionally (16). Depression showed a distinct profile: it was directly associated with hippocampal atrophy (19), and its direct associations with cognition remained significant after accounting for brain pathology. Depression also demonstrated significant total indirect effects on all cognitive outcomes via the combined brain mediators, consistent with partial mediation and suggesting contributions from both structural pathways and additional mechanisms not captured by the MRI mediators (18, 19). Overall, these findings indicate that modifiable risk factors relate to cognition through partially overlapping but mechanistically distinct pathways (55, 56).

An unexpected association emerged for obesity. Obesity was associated with better global cognitive status, lower odds of hippocampal atrophy, and slightly better performance on delayed recall and semantic fluency. Although this pattern may appear counterintuitive, similar findings have been reported in prior aging research. In late-life samples, individuals with more advanced neurodegenerative disease often experience unintentional weight loss or improved vascular indicators due to treatment or caregiver involvement (57,58). Cross-sectional analyses may therefore yield seemingly protective associations that reflect disease stage and survival effects rather than true benefit. Moreover, late-life obesity may not reflect midlife adiposity, which is more consistently associated with increased dementia risk, underscoring the need for longitudinal data to clarify the timing and clinical trajectory of obesity in relation to cognitive decline.

Beyond research implications, these findings have meaningful relevance for clinical practice and public health efforts aimed at reducing dementia risk (59–61). The additive pattern observed in this study suggests that interventions addressing multiple risk factors simultaneously may offer greater benefit than approaches that target one factor in isolation. This perspective is consistent with evidence from large multidomain prevention trials, including the Finnish Geriatric Intervention Study to Prevent Cognitive Impairment and Disability (FINGER) (59–61), the Multidomain Alzheimer Preventive Trial (MAPT) (61), and the Prevention of Dementia by Intensive Vascular Care (preDIVA) (61). Across these trials, multidomain programs integrating lifestyle, vascular, cognitive, and behavioral components have been shown to slow cognitive decline or maintain cognitive function in at-risk older adults (59–61). Although the specific interventions and outcomes differ across these studies, they collectively demonstrate that complex age-related cognitive conditions are more effectively addressed when several risk pathways are targeted together rather than separately.

This multidomain framework aligns directly with the results of the present study. The robust associations between cumulative risk burden and each brain and cognitive outcome indicate that broad, co-occurring patterns of risk may be driving much of the variability in late-life brain health. The observation that even small increases in total risk burden corresponded to widespread differences in WMHs, infarcts, hippocampal atrophy, global cognitive status, daily function, and memory performance reinforces the value of identifying individuals with clusters of modifiable risks. Clinically, such individuals may represent those most likely to benefit from multidomain prevention programs modeled on trials like FINGER, MAPT, and preDIVA. From a public health perspective, this suggests that strategies integrating vascular management, physical activity, nutrition, cognitive engagement, sensory health, and behavioral support may offer a more effective path toward reducing dementia risk at the population level than interventions that focus on only one risk at a time.

The findings also underscore the broader role of sensory, psychiatric, and social factors in shaping late-life cognition. The particularly strong associations for depression and low education point to pathways that reach beyond vascular mechanisms, highlighting the influence of mental health and lifelong learning experiences on cognitive resilience. These results are consistent with a large body of epidemiological work showing that both depression (62,63) and lower educational attainment (64–67) meaningfully increase dementia risk. Efforts that improve access to mental health care, support effective treatment of late-life depression, and expand opportunities for education and cognitive engagement across the lifespan may therefore play an important role in promoting healthier cognitive aging (68, 69).

Despite its strengths, the study has a few limitations. The cross-sectional design restricts inferences about the temporal ordering of risk exposure, brain changes, and cognitive decline. Given that timing, duration, and chronicity were not available for all risk factors, analyses could neither distinguish between long-standing exposure and more recent onset, nor could they address whether risk factors exert different effects at different disease stages. Participants in the NACC cohort are volunteers recruited through dementia research centers and tend to be more educated and healthier than the general population (70), which may limit generalizability and underestimate risk prevalence. Several risk factors relied on self-report or clinician documentation, and variables often lacked detail on treatment adherence, severity, or age of onset. MRI measures were based on qualitative ratings rather than volumetric quantification, which may reduce sensitivity to subtle structural changes in WMH volume or hippocampal integrity. Finally, although the sample included multiple racial groups, the majority of participants were White, limiting the ability to test whether associations vary across populations with different risk distributions, socioeconomic contexts, and structural health disparities.

Future research should address these limitations by incorporating longitudinal designs that track the evolution of risk factors, brain structure, and cognition over time. Such approaches would help clarify causal pathways, identify sensitive periods of risk exposure, and determine whether certain risk combinations accelerate decline more than others. Studies should also examine whether cumulative risk interacts with genetic factors, such as APOE e4, or social determinants, such as access to care. Additionally, future work should investigate whether multidomain interventions targeting risk clusters yield differential benefits depending on an individual’s baseline risk burden. Expanding analyses to more diverse populations is also essential, given established racial disparities in vascular burden, health care access, and dementia outcomes (71–74).

In conclusion, this study demonstrates that cumulative modifiable vascular, metabolic, sensory, psychiatric, and behavioral risk burden is strongly associated with structural brain injury and cognitive vulnerability in late life. Examining risk factors collectively provides a clearer and more comprehensive view of brain health than focusing on individual risks alone. Importantly, the observed associations with WMHs, cerebral infarcts, hippocampal atrophy, and multidomain cognitive decline represent changes that are well-established markers of heightened dementia risk. Thus, a greater cumulative modifiable risk burden may contribute to an increased likelihood of subsequent dementia diagnosis through its widespread impact on vascular injury, neurodegeneration, and cognitive impairment. These findings support multidomain prevention strategies and highlight the need for longitudinal, diverse, and mechanistically focused research to better understand the pathways through which modifiable risks shape late-life brain and cognitive outcomes.

## Data Availability

All data produced are available online at https://www.naccdata.org/

## Acknowledgments

The NACC database is funded by NIA/NIH Grant U24 AG072122. NACC data are contributed by the NIAfunded ADRCs: P30 AG062429 (PI James Brewer, MD, PhD), P30 AG066468 (PI Oscar Lopez, MD), P30 AG062421 (PI Bradley Hyman, MD, PhD), P30 AG066509 (PI Thomas Grabowski, MD), P30 AG066514 (PI Mary Sano, PhD), P30 AG066530 (PI Helena Chui, MD), P30 AG066507 (PI Marilyn Albert, PhD), P30 AG066444 (PI David Holtzman, MD), P30 AG066518 (PI Lisa Silbert, MD, MCR), P30 AG066512 (PI Thomas Wisniewski, MD), P30 AG066462 (PI Scott Small, MD), P30 AG072979 (PI David Wolk, MD), P30 AG072972 (PI Charles DeCarli, MD), P30 AG072976 (PI Andrew Saykin, PsyD), P30 AG072975 (PI Julie A. Schneider, MD, MS), P30 AG072978 (PI Ann McKee, MD), P30 AG072977 (PI Robert Vassar, PhD), P30 AG066519 (PI Frank LaFerla, PhD), P30 AG062677 (PI Ronald Petersen, MD, PhD), P30 AG079280 (PI Jessica Langbaum, PhD), P30 AG062422 (PI Gil Rabinovici, MD), P30 AG066511 (PI Allan Levey, MD, PhD), P30 AG072946 (PI Linda Van Eldik, PhD), P30 AG062715 (PI Sanjay Asthana, MD, FRCP), P30 AG072973 (PI Russell Swerdlow, MD), P30 AG066506 (PI Glenn Smith, PhD, ABPP), P30 AG066508 (PI Stephen Strittmatter, MD, PhD), P30 AG066515 (PI Victor Henderson, MD, MS), P30 AG072947 (PI Suzanne Craft, PhD), P30 AG072931 (PI Henry Paulson, MD, PhD), P30 AG066546 (PI Sudha Seshadri, MD), P30 AG086401 (PI Erik Roberson, MD, PhD), P30 AG086404 (PI Gary Rosenberg, MD), P20 AG068082 (PI Angela Jefferson, PhD), P30 AG072958 (PI Heather Whitson, MD), P30 AG072959 (PI James Leverenz, MD).

## Consent Statement

Written informed consent was obtained from participants or their study partner.

## Availability of data and materials

The data utilized in this study were also sourced from the National Alzheimer’s Coordinating Center (NACC) database (https://naccdata.org/), specifically drawing from the NACC Uniform Data Set (UDS) and MRI Data Set (Beekly et al., 2004; Besser, Kukull, Knopman, et al., 2018; Besser, Kukull, Teylan, et al., 2018).

---

**Table S1.** SEM Model Results: Indirect Associations Between Risk Factors and Cognitive Outcomes through each Brain Measure (c-paths)

|  | Infarcts |  |  |  | WMHs |  |  |  | Hippocampal Atrophy |  |  |  |
| --- | --- | --- | --- | --- | --- | --- | --- | --- | --- | --- | --- | --- |
| Risk Factor | Cognitive Status<br>(Std.all, *p*) | CDR-SB<br>(Std.all, *p*) | Delayed recall<br>(Std.all, *p*) | Semantic fluency<br>(Std.all, *p*) | Cognitive Status<br>(Std.all, *p*) | CDR-SB<br>(Std.all, *p*) | Delayed recall<br>(Std.all, *p*) | Semantic fluency<br>(Std.all, *p*) | Cognitive Status<br>(Std.all, *p*) | CDR-SB<br>(Std.all, *p*) | Delayed recall<br>(Std.all, *p*) | Semantic fluency<br>(Std.all, *p*) |
| Alcohol misuse | 0.00 (0.16) | 0.00 (0.38) | 0.00 (0.60) | 0.00 (0.85) | 0.00 (0.14) | 0.00 (0.52) | 0.00 (0.72) | 0.00 (0.17) | 0.00 (0.60) | -0.01 (0.60) | 0.01 (0.60) | 0.00 (0.60) |
| Smoking history | 0.00 (0.23) | 0.00 (0.41) | 0.00 (0.61) | 0.00 (0.85) | 0.00 (0.81) | 0.00 (0.82) | 0.00 (0.84) | 0.00 (0.81) | 0.00 (0.94) | 0.00 (0.94) | 0.00 (0.94) | 0.00 (0.94) |
| Hypercholesterolemia | 0.00 (0.20) | 0.00 (0.39) | 0.00 (0.60) | 0.00 (0.85) | 0.00 (0.33) | 0.00 (0.57) | 0.00 (0.73) | 0.00 (0.35) | 0.00 (0.44) | 0.01 (0.44) | -0.01 (0.44) | -0.01 (0.44) |
| Hypertension | 0.01 (0.08) | 0.00 (0.34) | 0.00 (0.59) | 0.00 (0.85) | 0.01 (0.05) | 0.00 (0.50) | 0.00 (0.72) | -0.01 (0.08) | 0.01 (0.05) | 0.02 (0.05) | -0.02 (0.05) | -0.01 (0.05) |
| Diabetes | 0.00 (0.57) | 0.00 (0.61) | 0.00 (0.70) | 0.00 (0.85) | 0.00 (0.98) | 0.00 (0.98) | 0.00 (0.98) | 0.00 (0.98) | 0.01 (0.36) | 0.01 (0.36) | -0.01 (0.36) | -0.01 (0.36) |
| Depression | 0.00 (0.27) | 0.00 (0.41) | 0.00 (0.61) | 0.00 (0.85) | 0.00 (0.68) | 0.00 (0.72) | 0.00 (0.78) | 0.00 (0.69) | 0.02 (**<.001**) | 0.05 (**<.001**) | -0.04 (**<.001**) | -0.04 (**<.001**) |
| Hearing loss | 0.00 (0.19) | 0.00 (0.40) | 0.00 (0.60) | 0.00 (0.85) | 0.00 (0.52) | 0.00 (0.64) | 0.00 (0.74) | 0.00 (0.53) | -0.01 (0.14) | -0.02 (0.14) | 0.01 (0.14) | 0.01 (0.14) |
| Vision loss | 0.00 (0.39) | 0.00 (0.49) | 0.00 (0.64) | 0.00 (0.85) | 0.00 (0.80) | 0.00 (0.81) | 0.00 (0.83) | 0.00 (0.80) | 0.01 (0.32) | 0.01 (0.32) | -0.01 (0.32) | -0.01 (0.32) |
| Obesity | 0.00 (0.81) | 0.00 (0.82) | 0.00 (0.82) | 0.00 (0.88) | 0.00 (0.57) | 0.00 (0.66) | 0.00 (0.75) | 0.00 (0.57) | -0.02 (**<.001**) | 0.04 (**<.001**) | 0.04 (**<.001**) | 0.03 (**<.001**) |
| Low education | 0.00 (0.59) | 0.00 (0.63) | 0.00 (0.70) | 0.00 (0.85) | 0.00 (0.53) | 0.00 (0.63) | 0.00 (0.75) | 0.00 (0.53) | 0.00 (0.79) | 0.00 (0.79) | 0.00 (0.79) | 0.00 (0.79) |

WMH = White Matter Hyperintensity. CDR-SB = Global Clinical Severity. Bolded values represent statistically significant *p*-values.

